# Patient-Reported Outcomes Correlate with Clinical Outcomes in Patients with Idiopathic Pulmonary Fibrosis

**DOI:** 10.1101/2025.05.27.25328368

**Authors:** Soumik Purkayastha, Suha Kadura, Cathie Spino, Ganesh Raghu

**Affiliations:** Department of Biostatistics and Health Data Science, University of Pittsburgh, Pittsburgh, PA, USA; Center for Health Equity Research and Promotion, VA Pittsburgh Healthcare System, US Department of Veterans Affairs, Pittsburgh, PA, USA; Division of Pulmonary, Critical Care and Sleep Medicine, Center for Interstitial Lung Diseases, Department of Medicine, University of Washington, Seattle, WA, USA; Department of Biostatistics, University of Michigan, Ann Arbor, MI, USA

## Abstract

**Background:** Idiopathic pulmonary fibrosis (IPF) is characterized by progressive fibrosis and declining quality of life. Patient-reported outcomes (PROs) provide a comprehensive understanding of a patient’s experience and health status than clinical measures (COs) alone, capturing how patients feel, function, and experience their health. However, the relationship between PROs and COs remains incompletely understood in this population.

**Methods:** We performed a secondary analysis of data from the YES-IPF trial, examining 60 IPF patients who completed a 12-week modified yoga program or continued usual activities. Pearson correlations between 21 PROs and 6 COs (FVC: absolute and percent-predicted, DLCO: absolute and percent-predicted, 6-minute walk test, and GAP: Gender-Age-Physiology index) were assessed at baseline and week. p-values were adjusted using the Benjamini-Hochberg method across 21 PROs for each CO measured.

**Findings:** Several significant correlations were observed between PROs and COs; key findings include <u>total L-IPF</u> correlating with <u>absolute FVC</u> having baseline *r* = −0.33 (*p* = 0.03) and 12-week *r* = 0.42 (*p* < 0.01), <u>total L-IPF</u> correlating with <u>6MWT</u> having baseline *r* = −0.43 (*p* < 0.01) and 12-week *r* = −0.37 (*p* = 0.02), <u>shortness of breath R-scale-PF</u> correlating with <u>absolute DLCO</u> having baseline*r* = −0.35 (*p* = 0.04) and 12-week *r* = −0.49 (*p* < 0.01), <u>total KBILD</u> correlating with <u>percent-predicted FVC</u> having baseline *r* = 0.38 (*p* = 0.03) and 12-week *r* = 0.43 (*p* < 0.01), and <u>K-BILD Breathlessness and Activities</u> score correlating with <u>GAP</u> score having baseline *r* = −0.42 (*p* = 0.02) and 12-week *r* = −0.40 (*p* = 0.02).

**Interpretation:** In IPF patients, breathlessness-specific PROs report correlation with COs of disease severity, while sleep-related and psychological domains demonstrate weaker correlations. This highlights that PROs and COs measure related but distinct aspects of IPF.

**Funding:** No funding support was received for this study.

**Trial Registration:** Registered with ClinicalTrials.gov (NCT0284862)

## MAIN TEXT

Idiopathic pulmonary fibrosis is a progressive lung disease significantly impacting patients’ quality of life. Patient-reported outcomes are increasingly recognized for their ability to capture patient experience of health, encompassing how they feel and function, which may not be reflected by clinical outcomes alone ^1^. Understanding the physical, emotional, and social impacts of patients with idiopathic pulmonary fibrosis, including the well-established burdens of dyspnea and cough, and the increasing recognition of fatigue, is paramount ^1,2^. While both patient-reported and clinical outcomes are valuable, patient-reported outcomes have not correlated with disease progression, defined as decline in forced vital capacity in patients with idiopathic pulmonary fibrosis in clinical trials to date.

The <u>Y</u>oga <u>E</u>ffect on Quality-of-Life <u>S</u>tudy Among Patients with <u>I</u>diopathic <u>P</u>ulmonary <u>F</u>ibrosis (YES-IPF) trial demonstrated that a 12-week modified yoga program improved patient-reported outcomes in patients with idiopathic pulmonary fibrosis, without concurrent significant changes in clinical outcomes like forced vital capacity ^3^. This prompted us to explore baseline as well as week 12 correlations for all patient-reported and clinical outcomes measured in the trial.

We analyzed data from 60 participants with well-defined idiopathic pulmonary fibrosis in accordance with the established criteria for diagnosis ^4^ who participated in the YES-IPF trial (<u>ClinicalTrials.gov: NCT02848625</u>). We pooled participants from the modified yoga and control group for all analyses. Descriptive statistics for these outcomes are presented in Table 1. Next, we computed pairwise Pearson correlations between 21 patient-reported outcomes (derived from seven validated instruments: the *Living with Idiopathic Pulmonary Fibrosis (L-IPF)* questionnaire ^2,3,5^, *Raghu-Scale for Pulmonary Fibrosis (R-scale-PF)* questionnaire ^3,6^, *King’s Brief Interstitial Lung Disease (KBILD)* questionnaire ^3,7^, *EuroQol-5D-5L, Hospital Anxiety and Depression Scale (HADS)* ^3^, *Patient-Reported Outcomes Measurement Information System (PROMIS)*: *Sleep Disturbance (SD)* and *Sleep-Related Impairment (SRI)*^8^ and *Epworth Sleepiness Scale (ESS)*^9^) and 6 clinical outcomes (*Forced Vital Capacity (FVC)*: *absolute* and *percent-predicted, Diffusing Capacity of the Lung for Carbon Monoxide (DLCO)*: *absolute* and *percent-predicted, 6-minute walk test (6MWT) distance, and the Gender-Age-Physiology (GAP) index*). While the first five clinical outcomes were measured during the trial, we also considered the GAP for this analysis. The GAP index is a validated prognostic staging system for idiopathic pulmonary fibrosis patients. Correlations were tested for significance cross-sectionally at baseline and at week 12. We performed complete case analyses for each correlation, given the extent of missingness was limited (ranging from 0 to 4). To control for multiple comparisons while maintaining the exploratory nature of this pilot study, p-values were adjusted using the Benjamini-Hochberg procedure ^10^ to control false discovery rate. Our findings are presented in Table 2. All analyses used R (version 4.4.1).

**Table 1.**
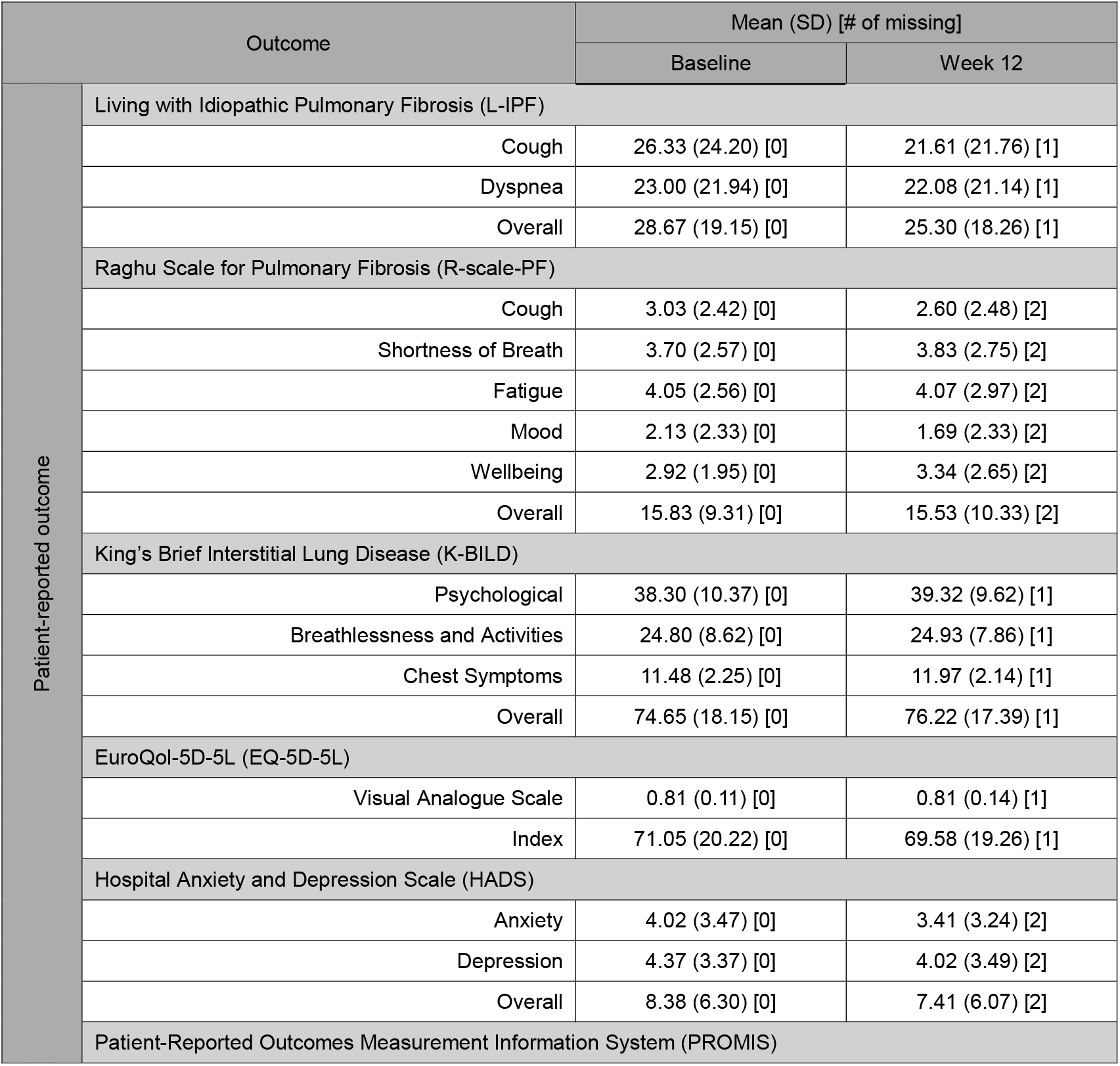
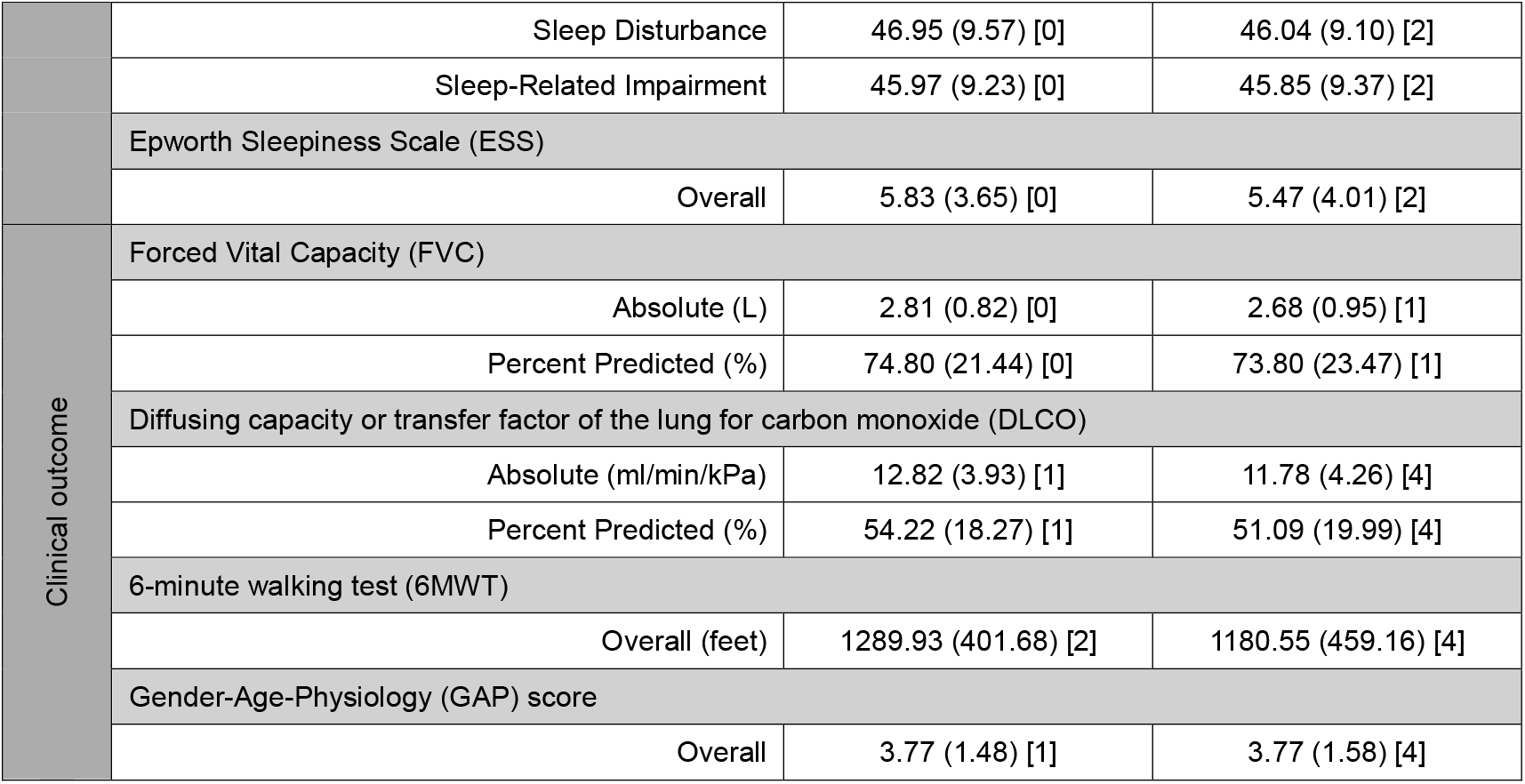
Mean (SD) [Count of Missing Measurements] of patient-reported and clinical outcomes at baseline and week 12 in the YES-IPF trial (N=60). Lower scores indicate better symptoms/quality of life for all outcomes except EuroQol-5D-5L scale, King’s Brief Interstitial Lung Disease scale, and the 6-minute walking test, where higher scores indicate better status.

**Table 2.** Pearson correlation coefficients (p-values) between patient-reported outcomes and clinical at baseline and week 12 in patients with idiopathic pulmonary fibrosis. P-values are adjusted using the Benjamini-Hochberg method to control for false discovery rate across 21 patient-reported outcomes for each of the 6 clinical outcomes. * Indicates statistical significance at adjusted p<0.05.

|  | Forced Vital Capacity (FVC) |  |  |  | Diffusing capacity or transfer factor of the lung for carbon monoxide (DLCO) |  |  |  | 6-minute walking test (6MWT) (feet) |  | Gender-Age-Physiology (GAP) score |  |
| --- | --- | --- | --- | --- | --- | --- | --- | --- | --- | --- | --- | --- |
|  | Absolute (L) |  | Pct. Predicted (%) |  | Absolute (L) |  | Pct. Predicted (%) |  | Baseline | Week 12 | Baseline | Week 12 |
|  | Baseline | Week 12 | Baseline | Week 12 | Baseline | Week 12 | Baseline | Week 12 |  |  |  |  |
| Living with Idiopathic Pulmonary Fibrosis (L-IPF) |  |  |  |  |  |  |  |  |  |  |  |  |
| Cough | -0.25<br>(0.10) | -0.26<br>(0.07) | -0.30<br>(0.11) | -0.30<br>(0.04) * | -0.21<br>(0.18) | -0.19<br>(0.23) | -0.21<br>(0.18) | -0.15<br>(0.45) | -0.21<br>(0.17) | -0.13<br>(0.44) | 0.22<br>(0.25) | 0.13<br>(0.66) |
| Dyspnea | -0.36<br>(0.03) * | -0.53<br>($< 0.01$) * | -0.28<br>(0.11) | -0.57<br>($< 0.01$) * | -0.36<br>(0.04) * | -0.51<br>($< 0.01$) * | -0.26<br>(0.18) | -0.35<br>(0.10) | -0.52<br>($< 0.01$) * | -0.56<br>($< 0.01$) * | 0.31<br>(0.13) | 0.47<br>($< 0.01$) * |
| Overall | -0.33<br>(0.03) * | -0.42<br>($< 0.01$) * | -0.25<br>(0.13) | -0.46<br>($< 0.01$) * | -0.31<br>(0.07) | -0.33<br>(0.04) * | -0.23<br>(0.18) | -0.18<br>(0.35) | -0.43<br>($< 0.01$) * | -0.37<br>(0.02) * | 0.22<br>(0.25) | 0.29<br>(0.12) |
| Raghu Scale for Pulmonary Fibrosis (R-scale-PF) |  |  |  |  |  |  |  |  |  |  |  |  |
| Cough | -0.09<br>(0.62) | -0.15<br>(0.30) | -0.25<br>(0.13) | -0.15<br>(0.35) | -0.18<br>(0.25) | -0.05<br>(0.78) | -0.25<br>(0.18) | -0.04<br>(0.80) | -0.06<br>(0.74) | 0.09<br>(0.55) | 0.23<br>(0.25) | -0.03<br>(0.89) |
| Shortness of Breath | -0.33<br>(0.03) * | -0.49<br>($< 0.01$) * | -0.32<br>(0.08) | -0.46<br>($< 0.01$) * | -0.35<br>(0.04) * | -0.49<br>($< 0.01$) * | -0.28<br>(0.18) | -0.34<br>(0.10) | -0.47<br>($< 0.01$) * | -0.43<br>($< 0.01$) * | 0.30<br>(0.13) | 0.36<br>(0.04) * |
| Fatigue | -0.14<br>(0.43) | -0.31<br>(0.04) * | -0.09<br>(0.62) | -0.21<br>(0.19) | -0.24<br>(0.13) | -0.34<br>(0.03) * | -0.22<br>(0.18) | -0.13<br>(0.47) | -0.36<br>(0.02) * | -0.31<br>(0.06) | 0.12<br>(0.48) | 0.10<br>(0.69) |
| Mood | -0.12<br>(0.50) | -0.19<br>(0.20) | 0.05<br>(0.74) | -0.04<br>(0.78) | -0.07<br>(0.67) | -0.15<br>(0.35) | 0.06<br>(0.68) | -0.04<br>(0.79) | -0.13<br>(0.42) | -0.12<br>(0.47) | -0.18<br>(0.34) | -0.08<br>(0.69) |
| Wellbeing | -0.33<br>(0.03) * | -0.35<br>(0.02) * | -0.21<br>(0.20) | -0.35<br>(0.02) * | -0.25<br>(0.13) | -0.31<br>(0.05) | -0.20<br>(0.20) | -0.25<br>(0.26) | -0.26<br>(0.10) | -0.30<br>(0.06) | 0.18<br>(0.34) | 0.15<br>(0.61) |
| Overall | -0.25<br>(0.10) | -0.38<br>($< 0.01$) * | -0.21<br>(0.20) | -0.32<br>(0.03) * | -0.28<br>(0.09) | -0.35<br>(0.03) * | -0.23<br>(0.18) | -0.21<br>(0.31) | -0.33<br>(0.03) * | -0.29<br>(0.07) | 0.17<br>(0.34) | 0.14<br>(0.63) |
| King's Brief Interstitial Lung Disease (K-BILD) |  |  |  |  |  |  |  |  |  |  |  |  |
| *Psychological* | 0.26<br>(0.10) | 0.29<br>(0.05) | 0.29<br>(0.11) | 0.28<br>(0.06) | 0.14<br>(0.41) | 0.24<br>(0.13) | 0.07<br>(0.68) | 0.18<br>(0.35) | 0.25<br>(0.10) | 0.18<br>(0.26) | -0.10<br>(0.57) | -0.09<br>(0.69) |
| *Breathlessness*<br>*and Activities* | 0.35<br>(0.03) * | 0.44<br>($< 0.01$) * | 0.39<br>(0.03) * | 0.51<br>($< 0.01$) * | 0.42<br>(0.02) * | 0.43<br>($< 0.01$) * | 0.31<br>(0.18) | 0.33<br>(0.10) | 0.54<br>($< 0.01$) * | 0.44<br>($< 0.01$) * | -0.42<br>(0.02) * | -0.40<br>(0.02) * |
| *Chest*<br>*Symptoms* | 0.14<br>(0.43) | 0.26<br>(0.07) | 0.20<br>(0.22) | 0.37<br>(0.01) * | 0.23<br>(0.15) | 0.17<br>(0.31) | 0.24<br>(0.18) | 0.13<br>(0.47) | 0.13<br>(0.42) | 0.19<br>(0.26) | -0.13<br>(0.46) | -0.09<br>(0.69) |
| *Overall* | 0.34<br>(0.03) * | 0.39<br>($< 0.01$) * | 0.38<br>(0.03) * | 0.43<br>($< 0.01$) * | 0.31<br>(0.07) | 0.35<br>(0.03) * | 0.22<br>(0.18) | 0.26<br>(0.26) | 0.42<br>($< 0.01$) * | 0.32<br>(0.05) | -0.27<br>(0.18) | -0.24<br>(0.24) |
| EuroQol-5D-5L (EQ-5D-5L) |  |  |  |  |  |  |  |  |  |  |  |  |
| *Visual Analogue*<br>*Scale* | 0.28<br>(0.09) | 0.35<br>(0.02) * | 0.26<br>(0.13) | 0.36<br>(0.02) * | 0.25<br>(0.13) | 0.33<br>(0.03) * | 0.22<br>(0.18) | 0.21<br>(0.31) | 0.33<br>(0.03) * | 0.44<br>($< 0.01$) * | -0.26<br>(0.19) | -0.29<br>(0.12) |
| *Index* | 0.34<br>(0.03) * | 0.38<br>($< 0.01$) * | 0.19<br>(0.25) | 0.39<br>($< 0.01$) * | 0.30<br>(0.07) | 0.34<br>(0.03) * | 0.16<br>(0.31) | 0.24<br>(0.26) | 0.49<br>($< 0.01$) * | 0.46<br>($< 0.01$) * | -0.11<br>(0.51) | -0.16<br>(0.56) |
| Hospital Anxiety and Depression Scale (HADS) |  |  |  |  |  |  |  |  |  |  |  |  |
| *Anxiety* | -0.05<br>(0.75) | -0.35<br>(0.02) * | 0.09<br>(0.62) | -0.20<br>(0.22) | 0.09<br>(0.67) | -0.15<br>(0.36) | 0.22<br>(0.18) | -0.05<br>(0.79) | -0.09<br>(0.63) | -0.19<br>(0.26) | -0.20<br>(0.30) | -0.02<br>(0.90) |
| *Depression* | -0.17<br>(0.34) | -0.15<br>(0.30) | 0.01<br>(0.95) | -0.10<br>(0.52) | -0.22<br>(0.16) | -0.27<br>(0.09) | -0.10<br>(0.54) | -0.18<br>(0.35) | -0.32<br>(0.03) * | -0.30<br>(0.06) | 0.07<br>(0.69) | 0.08<br>(0.69) |
| *Overall* | -0.12<br>(0.50) | -0.28<br>(0.06) | 0.05<br>(0.74) | -0.16<br>(0.32) | -0.07<br>(0.67) | -0.24<br>(0.14) | 0.07<br>(0.68) | -0.13<br>(0.47) | -0.22<br>(0.16) | -0.28<br>(0.08) | -0.07<br>(0.67) | 0.04<br>(0.89) |
| Patient-Reported Outcomes Measurement Information System (PROMIS) |  |  |  |  |  |  |  |  |  |  |  |  |
| *Sleep*<br>*Disturbance* | -0.07<br>(0.70) | -0.11<br>(0.43) | -0.09<br>(0.62) | -0.00<br>(0.99) | -0.01<br>(0.93) | -0.01<br>(0.94) | -0.06<br>(0.68) | 0.04<br>(0.79) | 0.03<br>(0.90) | 0.08<br>(0.60) | -0.05<br>(0.75) | -0.18<br>(0.50) |
| *Sleep-Related*<br>*Impairment* | -0.02<br>(0.91) | -0.22<br>(0.13) | 0.10<br>(0.62) | -0.11<br>(0.50) | 0.08<br>(0.67) | -0.04<br>(0.81) | 0.16<br>(0.31) | 0.09<br>(0.64) | -0.01<br>(0.95) | -0.10<br>(0.55) | -0.13<br>(0.46) | -0.11<br>(0.69) |
| Epworth Sleepiness Scale (ESS) |  |  |  |  |  |  |  |  |  |  |  |  |
| *Overall* | 0.02<br>(0.91) | -0.09<br>(0.51) | -0.05<br>(0.74) | -0.11<br>(0.50) | 0.03<br>(0.84) | 0.11<br>(0.50) | 0.11<br>(0.54) | 0.24<br>(0.26) | -0.01<br>(0.95) | 0.06<br>(0.66) | 0.02<br>(0.89) | -0.03<br>(0.89) |

We observed at baseline, patients demonstrated moderate disease burden with mean predicted FVC of 74.8% (SD 21.44), predicted DLCO of 54.2% (SD 18.27), and 6MWT distance of 1290 feet (SD 401.7). L-IPF cough and dyspnea reported mean scores of 26.3 (SD 24.2) and 23.0 (SD 21.9), respectively. K-BILD overall mean was 74.7 (SD 18.2), while HADS overall mean was 8.4 (SD 6.3).

We observed several statistically significant correlations (with false discovery rate < = 0.05) at both baseline and week 12. Quality of life measures of dyspnea and overall disease impact reported by patients showed consistent correlations with clinical measures: the L-IPF total score was negatively correlated with absolute FVC at baseline (*r* = −0.33, adjusted *p*= 0.03) and at week 12 (*r* = −0.42, adjusted *p*< 0.01). Similarly, the L-IPF total score was negatively correlated with 6MWT distance at baseline (*r* = −0.43, adjusted *p*< 0.01) and week 12 (*r* = −0.37, adjusted *p* = 0.02). Breathlessness-specific patient-reported outcomes also correlated significantly with FVC, DLCO, 6MWT, and GAP at one or both time points. For example, the R-scale-PF shortness of breath score showed negative correlations with absolute DLCO at baseline (*r* = −0.35, adjusted *p* = 0.04) and more strongly at week 12 (*r* = −0.49, adjusted p < 0.01). The K-BILD Breathlessness and Activities score negatively correlated with GAP score at baseline (*r* = −0.42, adjusted *p*= 0.02) and week 12 (*r* = −0.40, adjusted *p* = 0.02). The K-BILD total score, where higher scores indicate better health status, positively correlated with percent-predicted FVC at baseline (*r* = 0.38, adjusted p = 0.03) and week 12 (*r* = 0.43, adjusted p< 0.01). Finally, instruments assessing psychological aspects (HADS), and sleep (PROMIS and ESS) generally showed weaker or non-significant correlations with the clinical outcomes measured after false discovery rate adjustment.

Our analysis underscores that certain patient-reported outcomes, particularly those focused on respiratory symptoms like dyspnea, correlate with objective clinical outcomes of lung function and exercise capacity and further capture distinct aspects of the patient experience. The moderate strength of most observed correlations, even after rigorous adjustment for multiple comparisons, suggests that patient-reported outcomes provide unique, complementary information to clinical outcomes, rather than merely reflecting the same constructs. This aligns with previous research suggesting that patient-reported outcomes and clinical outcomes can offer different perspectives on a patient’s condition ^1^. The application of the Benjamini-Hochberg procedure allowed us to identify these relationships with controlled false discovery rate, which is particularly suitable for hypothesis-generating research in pilot studies. The weaker correlations observed for psychological and sleep-related patient-reported outcomes with physiological measures highlight the multifaceted impact of idiopathic pulmonary fibrosis, where these domains may be influenced by factors beyond those captured by standard lung function tests. We acknowledge limitations for our study findings: small sample size, single center study, and a short duration of follow up at only 12 weeks; these issues may limit power to detect weaker correlations.

In conclusion, our analysis demonstrates that, in patients with idiopathic pulmonary fibrosis, patient-reported outcomes relating to breathlessness and overall disease impact show significant correlations with established clinical measures at baseline and follow up at 12 weeks. The distinct patterns and strengths of these correlations emphasize that patient-reported outcomes and clinical outcomes are not interchangeable and offer complementary insights. This work supports the continued integration of patient-reported outcomes alongside clinical outcomes in clinical trials and practice to gain a complete understanding of idiopathic pulmonary fibrosis. Given the observed correlation of patient-reported outcomes with conventional clinical measures in idiopathic pulmonary clinical trials and practice – a strong rationale exists for utilizing patient-reported outcomes as surrogate or primary endpoints in future studies to better capture patient-centric benefits.

## Data Availability

All data produced in the present study are available upon reasonable request to the authors.

